# COMBINATION OF TOCILIZUMAB AND STEROIDS TO IMPROVE MORTALITY IN PATIENTS WITH SEVERE COVID-19 INFECTION: A SPANISH, MULTICENTER, COHORT STUDY

**DOI:** 10.1101/2020.09.07.20189357

**Authors:** Belén Ruiz-Antorán, Aránzazu Sancho-López, Ferran Torres, Víctor Moreno-Torres Concha, Itziar de Pablo López de Abechuco, Paulina García López, Francisco Abad-Santos, Clara María Rosso Fernández, Ana Aldea-Perona, Eva Montané, Ruth M. Aparicio-Hdez, Roser LLop Rius, Consuelo Pedrós, Paloma Gijón, Carolina Hernández Carballo, María José Pedrosa Martínez, Consuelo Rodríguez Jiménez, Guillermo Prada-Ramallal, Lourdes Cabrera García, Josefa Andrea Aguilar García, Rocío Sanjuan-Jimenez, Evelyn Iveth Ortiz Barraza, Enrique Sánchez-Chica, Ana Fernández-Cruz, on behalf of the TOCICOV-study group

**Author notes:** Contributed equally (Belén Ruiz-Antorán, Aránzazu Sancho-López). Corresponding author Dr. Ana Fernández Cruz Infectious Diseases Unit Internal Medicine Department Hospital Universitario Puerta de Hierro-Majadahonda, Madrid, Spain.

## Abstract

**Background:** We aimed to determine the impact of tocilizumab use in severe COVID-19 pneumonia mortality.

**Methods:** We performed a multicentre retrospective cohort study in 18 tertiary hospitals in Spain, from March to April 2020. Consecutive patients admitted with severe COVID-19 treated with tocilizumab were compared to patients not treated with tocilizumab, adjusting by Inverse Probability of the Treatment Weights (IPTW). Tocilizumab effect in patients receiving steroids during the 48h following inclusion was analyzed.

**Results:** During the study period, 506 patients with severe COVID-19 fulfilled inclusion criteria. Among them, 268 were treated with tocilizumab and 238 patients were not. Median time to tocilizumab treatment from onset of symptoms was 11 days (IQR 8-14). Global mortality was 23.7%. Mortality was lower in patients treated with tocilizumab than in controls (16.8% versus 31.5%, HR 0.514 [95CI 0.355-0.744], p< 0.001; weighted HR 0.741 [95CI 0.619-0.887], p = 0.001). Tocilizumab treatment reduced mortality by 14.7% relative to no tocilizumab treatment (RRR 46.7%). We calculated a number necessary to treat of 7. Among patients treated with steroids, mortality was lower in patients treated with tocilizumab than in those treated with steroids alone (10.9% versus 40.2%, HR 0.511 [95CI 0.352-0.741], p = 0.036; weighted HR 0.6 [95CI 0.449-0.804], p< 0.001) (Interaction p = 0.094).

**Conclusions:** These results show that survival of patients with severe COVID-19 is higher in patients treated with tocilizumab than in those not treated, and that tocilizumab effect adds to that of steroids administered to non-intubated cases with COVID-19 during the first 48 hours of presenting with respiratory failure despite of oxygen therapy. Randomised controlled studies are needed to confirm these results.

**Summary:** We investigated in-hospital mortality of patients with severe SARS-CoV-2 pneumonia in a multicenter series of patients treated with tocilizumab compared to controls, and adjusted using IPTW. Our results show a beneficial impact of tocilizumab treatment in SARS-CoV-2 pneumonia, that adds to that of steroids.

## INTRODUCTION

In the past few decades, newly evolved Coronaviruses have posed a global threat to public health.

The most recent one, SARS-CoV-2 coronavirus, has produced a pandemic infecting more than 10 million people and causing more than 500 thousand deaths in the world, challenging not only healthcare systems but also the culture and economy of the population[1].

SARS-CoV-2 causes COVID-19 disease, which in most cases is an asymptomatic or mildly symptomatic condition that resolves without therapy or with minor supportive treatment. In some cases, SARS-CoV-2 can cause pneumonia which in 20% may be moderate in severity. However, a subgroup of COVID-19 patients with pneumonia develops rapidly progressing respiratory failure that may necessitate mechanical ventilation and support in an intensive care unit (ICU), and/or multi-organ and systemic manifestations in terms of sepsis, septic shock, and multiple organ dysfunction syndromes [2].

Available evidence suggests that a hyper-inflammatory syndrome (HIS) that resembles secondary hemophagocytic lymphohistiocytosis (sHLH) may have a pathogenic role [3,4].This is consistent with the general knowledge about coronavirus (CoV) infections, where immune response has been shown to play an essential role to control and eliminate CoV infections, and maladjusted immune responses may result in immunopathology and impaired pulmonary gas exchange[5].

At the time of conducting the study, treatment was mainly symptomatic, with oxygen therapy representing the major treatment intervention for patients with severe infection. Several antiviral treatments have been used such as lopinavir/ritonavir, chloroquine, hydroxychloroquine, remdesivir, and alpha-interferon. No vaccine is currently available. Based on the observation that acute, life-threatening respiratory injury associated with an over-exuberant cytokine release occurred in some patients infected by SARS-CoV-2, treatment strategies aimed to break down this hyperinflammatory response were considered. Elevated levels of blood interleukin-6 (IL-6) have been identified as a one of the risk factors associated with severe COVID-19 disease [5].

In this context, tocilizumab, a humanized IgG1 monoclonal antibody targeting the IL-6 receptor, was postulated as a suitable treatment option[6]. Preliminary experiences with tocilizumab showed promising results with improvements seen in clinical symptoms, oxygenation status, and inflammatory laboratory parameters[7]. This limited evidence together with a strong mechanistic rationale, plus current knowledge on the role of anti-IL6 inhibitors in the treatment of cytokine release syndromes, supported the progressively extended use of tocilizumab in clinical practice. Simultaneously numerous clinical studies, including clinical trials, were initiated around the world with the aim to assess the potential efficacy of tocilizumab in preventing the fatal consequences of acute respiratory and multi-organ failure associated with acute respiratory syndrome due to severe COVID-19 infection [8].

At the time of launching our study (3rd March 2020), many uncertainties existed on the actual benefits and risks of tocilizumab in the treatment of SARS-CoV-2 pneumonia. To increase knowledge on the actual role of tocilizumab in SARS-CoV-2 pneumonia, we conducted a retrospective cohort multicentre study. Our study was aimed to describe our experience with a series of consecutive COVID-19 patients treated with TCZ in 18 tertiary hospitals in Spain and compare the outcomes of this cohort with those observed in a similar cohort of SARS-CoV-2 infected patients who did not receive tocilizumab, to identify patients who will most benefit from its use.

## PATIENTS AND METHODS

### Design, study period and subjects

This a retrospective, observational cohort study performed in 18 tertiary Spanish hospitals, including patients with severe COVID infection

The study population were adult patients (≥18 years) with COVID-19, confirmed by PCR on nasopharyngeal swab, who were consecutively admitted to the participating hospitals between March 3th and April 20th.

Patients were included consecutively according to the date of admission to hospital until the planned sample size was met in a competitive manner. Eighteen centres contributed data on in both cohorts (Appendix Table 1). Eligible patients were hospitalized patients outside the Intensive Care Units with documented interstitial pneumonia with severe respiratory failure (respiratory severity scale BRESCIA-COVID = 2) (or rapid deterioration of respiratory exchanges without immediate possibility of invasive ventilation (respiratory severity scale COVID = 3) and at least one of these parameters: IL6 > 40 pg/mL, or increasing LDH or LDH> twice upper limit of normal, increasing CRP, D-Dimer (DD) > 1,500 ng / mL, Lymphocytes < 1200/µL or ferritin> 500 ng / ml. We excluded patients younger than 18 years and those who died within 24 hours after admission to the hospital or after developing inclusion criteria. Of these, patients who received tocilizumab therapy according to clinical practice were assigned to the tocilizumab cohort, whilst patients who did not were assigned to the control cohort.

### Data collection

Epidemiological, clinical, pharmacological, laboratory and radiologic data were extracted from medical records using a standardized data collection form. The patients were followed according to clinical practice. Data were collected from days 1, 3, 7, 15 and 28 post-inclusion. Adverse events possibly or probably related to tocilizumab treatment were collected for treated patients. All data were included by a primary reviewer and subsequently checked by two senior physicians.

### Laboratory procedures

Routine blood examinations included a complete blood count, coagulation profile, serum biochemical tests (including lactate dehydrogenase), C reactive protein (CRP), DD, interleukin-6 (IL-6), and serum ferritin. Chest radiographs or CT scans were also done for all inpatients.

### Definition of the outcome

The primary outcome of the study was in-hospital mortality. The outcome of patients treated with tocilizumab was compared to that of those who did not receive tocilizumab. In patients from the Non-treated cohort, baseline day (day 0) was defined as the first day that the patient fulfilled the inclusion criteria established in the protocol. In patients from the tocilizumab cohort, baseline day was considered the day of initiation of treatment with tocilizumab.

### Definition of the exposure

Exposure to tocilizumab was defined as the use of intravenous tocilizumab at any time during the hospital admission. The decision to prescribe tocilizumab was at the discretion of the treating physician.

Details of tocilizumab use (including the timing of initiation, dosing, and number of doses) were recorded. Likewise, the choice of COVID treatments other than tocilizumab was at the discretion of the treating physician, although based on national and local recommendations for COVID-19 management.

For the main analysis, we generated a variable with the following mutually exclusive categories: “non-use of tocilizumab drug” (control cohort) and “use of tocilizumab drug” (treatment cohort). Subsequently, for the treatment cohort, we disaggregated the latter into two different subgroups: subgroup with concomitant steroids and no concomitant corticosteroid group. Concomitant use of corticosteroids was defined as the initial use of corticosteroid within the first 48 hours after administration of tocilizumab (steroid-48h).

### Statistical analysis

Categorical variables are described with frequencies and percentages and continuous variables with mean (standard deviation -SD-) and median [interquartile range: 25th-75th percentiles], and the survival function is described using the Kaplan-Meier function.

We used standardized differences, defined as differences between groups divided by pooled standard deviation to assess heterogeneity between both cohorts for baseline covariates. The Inverse Probability of the Treatment Weights (IPTW) approach[9] was used to create a pseudo-population in which the 2 groups were balanced across baseline covariates. The stabilised weights were calculated using propensity scores (PS) [10] obtained from a logistic regression model aimed to minimise the between arms standardized differences[11]. The baseline covariates included in the final model were gender, age, hypertension, neurologic exploration, diabetes mellitus, WHO ordinal scale, time from symptoms, confirmed infection, lymphocytes, neutrophils, platelets, prothrombin activation, temperature, LDH, and baseline medication use of ACEs inhibitors, lopinavir-ritonavir, hydroxychloroquine, corticosteroids, interferon, non-steroidal anti-inflammatory drugs, moxifloxacin, remdesivir, azithromycin. Covariate balance was assessed using the standardised differences with the goal to achieve values < 0.10 using the IPTW to define insignificant difference in potential confounders, which achieved by most, but not by all baseline covariates. However, for some authors < 0.20 might be also acceptable[12, 13] and only interleukin 6 (IL-6) remained as unbalanced (STD = 0.42) with no possible PS correction given the high number of missing data (≈51%).

Since there were missing data for some key covariates needed for the propensity score calculations, the following variables (all with less of 10% missingness) were imputed using the EM algorithm which relies on the flexible and reasonable Missing At Random assumption: WHO ordinal scale, time from symptoms, lymphocytes, neutrophils, platelets, haemoglobin, PCR, D-dimer, temperature, LDH and prothrombin activation. Baseline categorical data were compared using the chi-square test and continuous variables using ANOVA with rank-transformed data, for raw and IPTW adjusted analyses. Raw and IPTW adjusted regression models were used to estimates risks (hazard ratio (HR) with 95%CI (95%CI)) for time to event variables.

In all statistical analyses, we applied a two-sided type I error of 5%. The software SPSS v.25 (IBM Corp., Armonk, NY) and SAS v9.4 (Cary, NC, USA) were used.

#### Ethics

The study was approved by Spanish Agency for Medicine and Health Product and by the Institutional Review Board (CEIm) at Hospital Universitario Puerta de Hierro-Majadahonda (FIB-TOC-2020-01), and a waiver for the informed consent was granted. The study complied with the provisions in European Union (EU) and Spanish legislation on data protection and the Declaration of Helsinki 2013.

Registration: The protocol of the study was registered in EU PAS Register #EUPAS34415 on 31th March 2020 and publicly available at: http://www.encepp.eu/encepp/viewResource.htm?id=35237

### RESULTS

During the study period, 506 consecutive patients with COVID-19 and respiratory insufficiency and increased inflammatory parameters were included. Among them, 268 were treated with tocilizumab and 238 patients were not. Twenty-four patients who died in the first 24h after developing inclusion criteria were excluded.

#### Baseline clinical characteristics

Characteristics of both cohorts are shown in table 1. Median time to tocilizumab from the onset of symptoms was 11 days (IQR 8-14 days).

**Table 1.** Baseline characteristic of patients with SARS-CoV-2 infection according to tocilizumab exposure

|  | Raw analysis |  |  |  | IPTW analysis |  |  |
| --- | --- | --- | --- | --- | --- | --- | --- |
|  | Tocilizumab group (N =268) | Control group (N =238) | p value | Standardised difference | Tocilizumab group (N =254) | Control group (N =235) | Standardised difference |
| Gender (Men), n (%) | 184 (68.7) | 140 (58.8) | 0,021 | -20.6% | 165 (65.0) | 150 (63.8) | 0.9% |
| Age, Mean (SD) | 65.0 (11.7) | 71.3 (14.2) | <.0001 | -47.9% | 66.6 (10.7) | 67.3 (14.8) | -1.8% |
| Underlying Medical Conditions, n (%) |  |  |  |  |  |  |  |
| High Blood pressure | 130 (48.5) | 145 (60.9) | 0,001 | -25.1% | 132 (52.0) | 126 (53.5) | -0.4% |
| Cardiovascular disease | 65 (24.3) | 74 (31.1) | 0,084 | -15.3% | 68 (26.9) | 61 (26.1) | 3.5% |
| Diabetes | 78 (29.1) | 68 (28.6) | 0,894 | 1.2% | 75 (29.6) | 66 (27.8) | 5.2% |
| Chronic kidney disease | 23 (8.6) | 30 (12.6) | 0,140 | -13.1% | 23 (9.2) | 23 (9.9) | -0.9% |
| Onco-hematologic | 12 (4.5) | 12 (5.0) | 0,765 | -2.7% | 10 (3.8) | 9 (4.0) | -0.1% |
| Chronic lung disease | 46 (17.2) | 49 (20.6) | 0,324 | -8.8% | 47 (18.4) | 43 (18.4) | -0.3% |
| Transplant (SOT/SCT) | 6 (2.2) | 4 (1.7) | 0,652 | 4.0% | 6 (2.2) | 7 (3.0) | -4.0% |
| Neurologic | 15 (5.6) | 42 (17.6) | <.0001 | -38.3% | 21 (8.2) | 25 (10.4) | -6.1% |
| Liver disease | 10 (3.7) | 12 (5.0) | 0,470 | -6.4% | 15 (5.9) | 13 (5.5) | 2.7% |
| HIV | 2 (0.7) | 3 (1.3) | 0,559 | -5.2% | 1 (0.5) | 2 (0.7) | -1.9% |
| NSAIDs | 19 (7.1) | 18 (7.6) | 0,838 | -1.8% | 22 (8.6) | 16 (6.7) | 8.5% |
| ACE inhibitors | 53 (19.8) | 40 (16.8) | 0,389 | 7.7% | 47 (18.4) | 41 (17.4) | 3.4% |
| ARBs | 49 (18.3) | 56 (23.5) | 0,146 | -12.9% | 61 (23.8) | 50 (21.0) | 8.9% |
| Treatment, n (%) |  |  |  |  |  |  |  |
| Hydroxychloroquine | 262 (97.8) | 220 (92.4) | 0,004 | 24.9% | 247 (97.1) | 223 (94.8) | 11.0% |
| Lopinavir/Ritonavir | 240 (89.6) | 164 (68.9) | <.0001 | 52.6% | 206 (81.0) | 180 (76.3) | 10.0% |
| Azithromycin | 163 (60.8) | 132 (55.5) | 0,222 | 10.9% | 150 (59.0) | 138 (58.4) | -2.5% |
| Remdesivir | 1 (0.4) | 1 (0.4) | 0,932 | -0.8% | 1 (0.4) | 1 (0.4) | 0.5% |
| Interferon | 124 (46.3) | 72 (30.3) | <.0001 | 33.4% | 105 (41.2) | 92 (39.0) | 1.3% |
| Steroids |  |  |  |  |  |  |  |
| - Steroids prior to D0 | 87 (32.5) | 26 (10.9) | <.0001 | 54.1% | 61 (23.8) | 51 (21.5) | -1.2% |
| WHO classification D0 |  |  |  |  |  |  |  |
| Admitted, no oxygen therapy | 5 (1.9) | 5 (2.1) | 0,981 |  | 4 (1.6) | 4 (1.6) |  |
| Admitted with oxygen therapy | 244 (91.0) | 216 (90.8) | 0,981 | 0.6% | 234 (91.9) | 217 (92.3) | 1.9% |
| High flow | 19 (7.1) | 17 (7.1) | 0,982 |  | 16 (6.5) | 14 (6.1) |  |
| Laboratory values, Mean (SD) |  |  |  |  |  |  |  |
| Lymphocytes (x10E3/mm<sup>3</sup>) | 834.3 (469.7) | 902.9 (580.9) | 0,235 | -13.0% | 843.3 (420.5) | 854.1 (501.8) | -0.6% |
| Lactate dehydrogenase (UI/L) | 451.4 (186.2) | 459.8 (339.8) | 0,361 | -3.1% | 448.8 (179.6) | 463.16 (288.2) | -9.1% |
| D Dimer (ng/ml) | 2,200.2 (5,216.8) | 1,950.9 (2,588.7) | 0,334 | 6.1% | 2,040.8 (4,412.3) | 1,869.9 (2,553.4) | -1.5% |
| C-reactive protein (mg/l) | 149.5 (105.7) | 148.0 (111.1) | 0,584 | 1.4% | 148.54 (95.64) | 146.0 (105.9) | 4.6% |
| Ferritin (ng/ml) | 1,682.6 (1,427.9) | 1,363.7 (2,953.5) | <.0001 | 13.7% | 1,697.1 (1,388.3) | 1,532.0 (2,794.2) | -0.6% |
| Interleukin 6 (IL-6) (pg/ml) | 206.5 (468.5) | 78.8 (94.9) | <.0001 | 37.8% | 195.4 (375.2) | 80.3 (93.7) | 42.4% |
| Chest CT (at hospital admission), n (%) |  |  |  |  |  |  |  |
| Normal imaging | 4 (1.6) | 9 (4.1) |  |  | 3 (1.3) | 7 (3.2) |  |
| Unilateral pneumonia | 18 (7.3) | 26 (11.9) |  |  | 17 (7.2) | 25 (12.3) |  |
| Bilateral interstitial pneumonia | 66 (26.9) | 58 (26.6) | 0.192 |  | 65 (27.7) | 54 (24.7) |  |
| Patchy bilateral pneumonia | 100 (40.8) | 77 (35.3) |  |  | 91 (38.7) | 80 (36.5) |  |
| Confluent bilateral pneumonia | 57 (23.3) | 48 (22.0) |  |  | 59 (38.7) | 61 (23.3) |  |
| Tocilizumab treatment, n (%) |  |  |  |  |  |  |  |
| Initial tocilizumab dose (mg) | 532.4 (109.5) |  |  |  | 538.8 (110.1) |  |  |
| Cumulative tocilizumab dose (mg) | 789.2 (354.8) |  |  |  | 827.9 (366.1) |  |  |
| Time from onset of symptoms to D0, Mean (SD) | 11.73 (5.20) | 8.43 (4.67) | <.0001 | 66.7% | 10.41 (4.71) | 10.12 (5.42) | -0.2% |

Among 268 patients treated with tocilizumab, 22 (8.2%) received 3 doses, 92 (34.3%) received 2 doses and the remaining 154 (57.4%) received only one dose. Median initial tocilizumab dose was 600 mg (IQR 400-600). Median time from first dose to second dose was 1 day (IQR 1-1), and from first dose to third dose was 2 days (IQR 1-3). Median cumulative dose of tocilizumab was 600 mg (IQR 600-1000).

A propensity score was developed to estimate each patient’s probability of receiving tocilizumab given their baseline characteristics and reduce selection bias. Characteristics of the treatment and control cohort after adjustment by IPTW (inverse probability treatment weighting) are shown in table 1.

#### In-hospital mortality of patients treated with tocilizumab compared to patients not treated with tocilizumab

Global hospital mortality was 23.7%. Characteristics of survivors and non-survivors are shown in Appendix Table 2.

Mortality was lower in patients treated with tocilizumab than in controls (16.8% versus 31.5%, HR 0.514 [95CI 0.355-0.744], p< 0.001) (Table 2). Tocilizumab treatment reduced mortality by 14.7% relative to no tocilizumab treatment (RRR 46.7%). We calculated a number necessary to treat of 7. Differences in mortality persisted after applying the propensity score adjusted for tocilizumab treatment (weighted HR 0.741 [95CI 0.619-0.887], p = 0.001) (Table 2). Figure 1 demonstrates differences in the probability for survival for patients with SARS-CoV2 infection, according to tocilizumab treatment (log-rank p< 0.001).

**Table 2.** Association between tocilizumab treatment and in-hospital mortality in patients with SARS-COV-2 infection, according to tocilizumab and steroid-48 exposure.

| Number of events |  |  | Ratio |  | IPTW ratio |  |
| --- | --- | --- | --- | --- | --- | --- |
| All | Tocilizumab<br>N =268 | No tocilizumab<br>N = 238 | HR<br>(95% CI) | p value | wHR<br>(95% CI) | p value |
| Deaths | 45 (16.8%) | 75 (31.5%) | 0.514<br>(0.355-0.744) | **<0.001** | 0.741<br>(0.619-0.887) | **0.001** |
| **Steroid-48** | **N=119** | **N=87** |  |  |  |  |
| Deaths | 13 (10.9%) | 35 (40.2%) | 0.511<br>(0.352-0.741) | **<0.001** | 0.600<br>(0.449-0.804) | **0.006** |
| **NO Steroid-48** | **N=149** | **N=151** |  |  |  |  |
| Deaths | 32 (21.5%) | 40 (26.5%) | 0.713<br>(0.447-1.137) | 0.345 | 0.830<br>(0.650-1.048) | 0.115 |

HR=hazard ratio; IPTW=inverse probability of treatment weighting; wHR=weighted hazard ratio; steroid-48: treated with steroids in the 48h following inclusion.
\*Weighted hazard ratios and 95% confidence intervals were obtained by inverse probability treatment weighting

**Figure 1.**
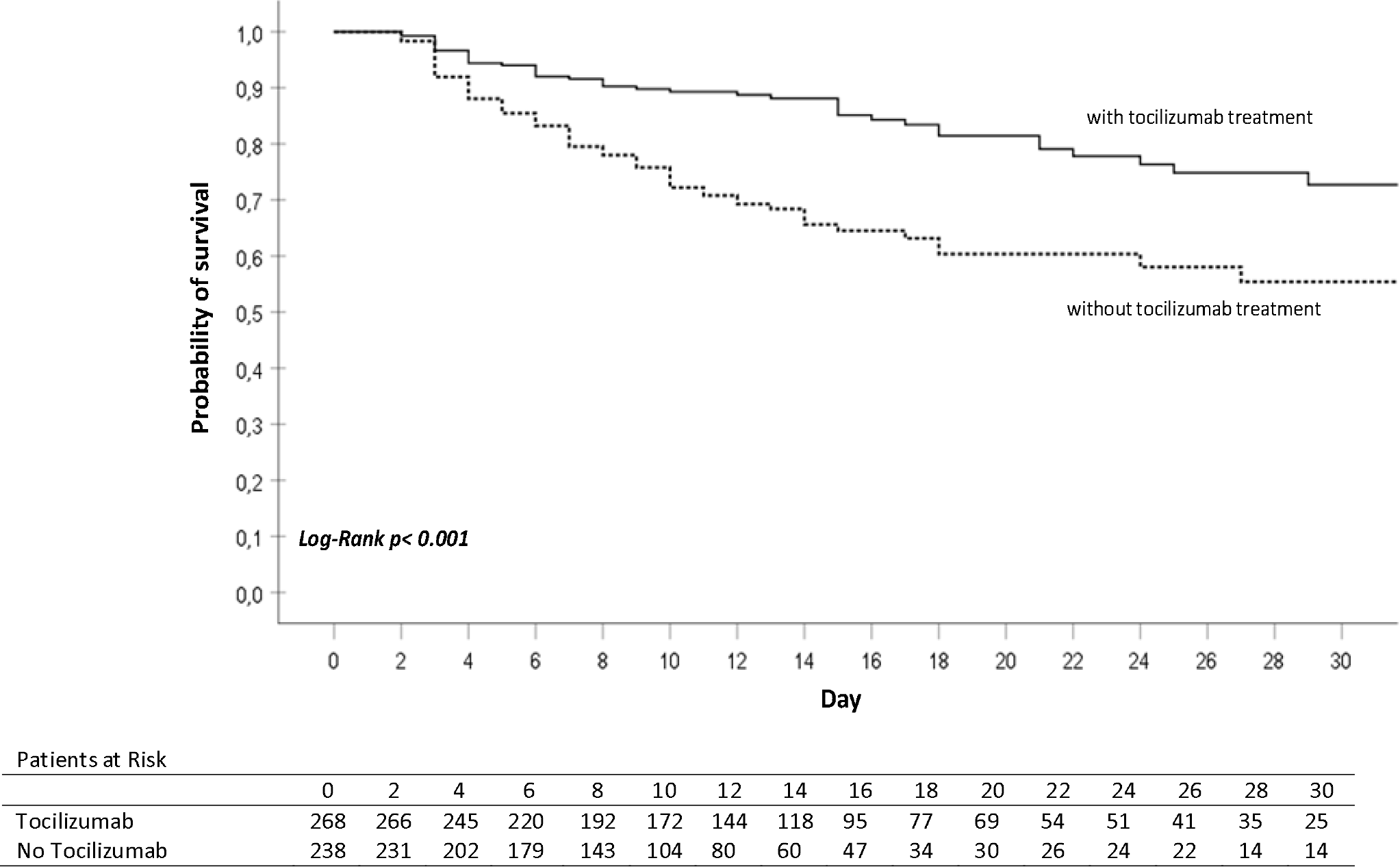
Probability of survival of patients with SARS-COV-2 infection, according to tocilizumab exposure. Descriptive raw analysis

#### Tocilizumab effect in subgroups according to baseline characteristics

The effect of tocilizumab treatment on mortality in different subsets of patients was consistent with a protective effect. Figure 2 shows different effect of tocilizumab according to different values of variables that displayed significant interaction. Patients older than 65 years, with lymphocyte counts below 1000 cells/μL, hypertension and cardiovascular disease seem to benefit the most from tocilizumab treatment.

**Figure 2.**
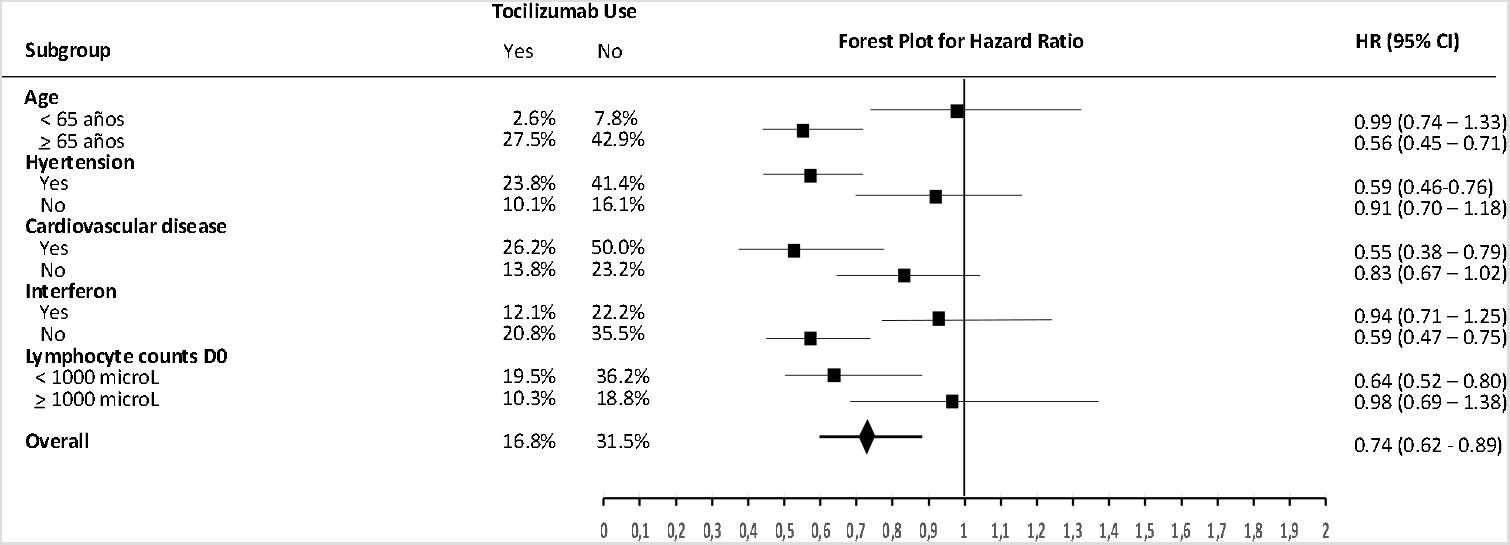
Forest plot of stratified analyses for mortality showing the weighted hazard ratio from IPTW analysis of tocilizumab treatment.

#### Effect of tocilizumab in patients with post-baseline steroid treatment

One-hundred nineteen (44.4%) patients in the tocilizumab arm and 87 (36.6%) in the control arm were treated with steroids in the 48h following inclusion (steroids-48). Among them 71.3% patients received steroid in pulses, and 28.7% non-pulsed steroids. Characteristics of patients according to tocilizumab and steroid-48 exposure can be seen in Appendix table 3.

An interaction was found between tocilizumab and steroid-48 use (interaction p = 0.094). Among patients treated with steroids-48, mortality was lower in patients treated with tocilizumab than in those treated with steroids-48h alone (10.9% versus 40.2%, HR 0.511 [95CI 0.352-0.741], p = 0.036; weighted HR 0.6 [95CI 0.449-0.804], p< 0.001) (Table 2).

Tocilizumab with steroid-48 treatment reduced mortality by 29.1% relative to no tocilizumab treatment (RRR 72.8%). We calculated a number necessary to treat of 4. Figure 3 shows differences in probability of survival at according to steroid exposure in patients treated with tocilizumab.

**Figure 3.**
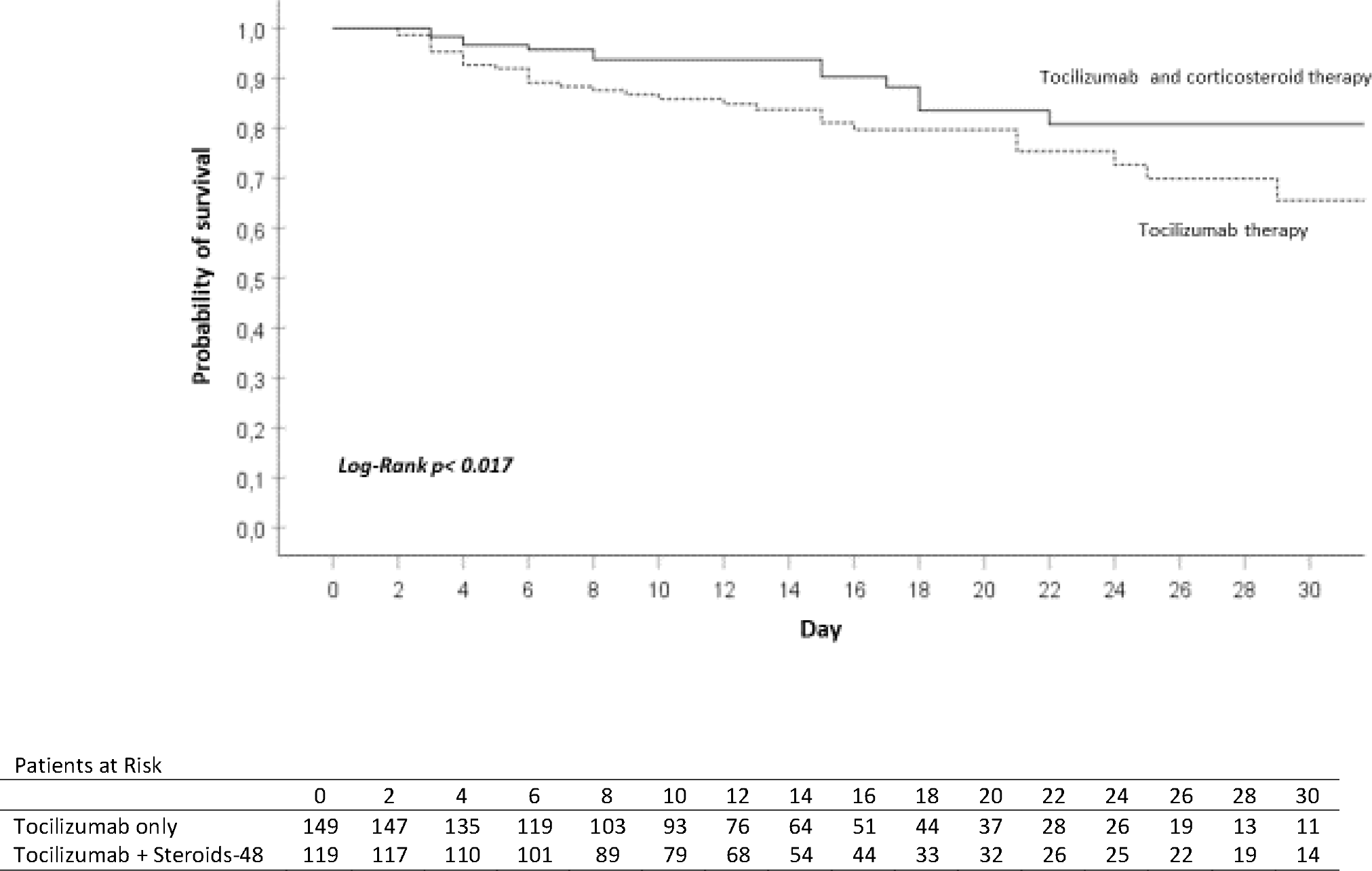
Probability of survival of patients with SARS-COV-2 infection treated with tocilizumab, according to steroid-48 exposure.

#### Variation over time of prognostic biomarkers

Figure 4 displays variation over time of C- reactive protein, IL-6, LDH, DD, lymphocyte counts and ferritin, according to tocilizumab exposure (figure 4A), and mortality in the subgroup of patients treated with tocilizumab (figure 4B). IL-6 levels were available only for 51 patients at baseline, and 68 patients during follow-up.

**Figure 4.**
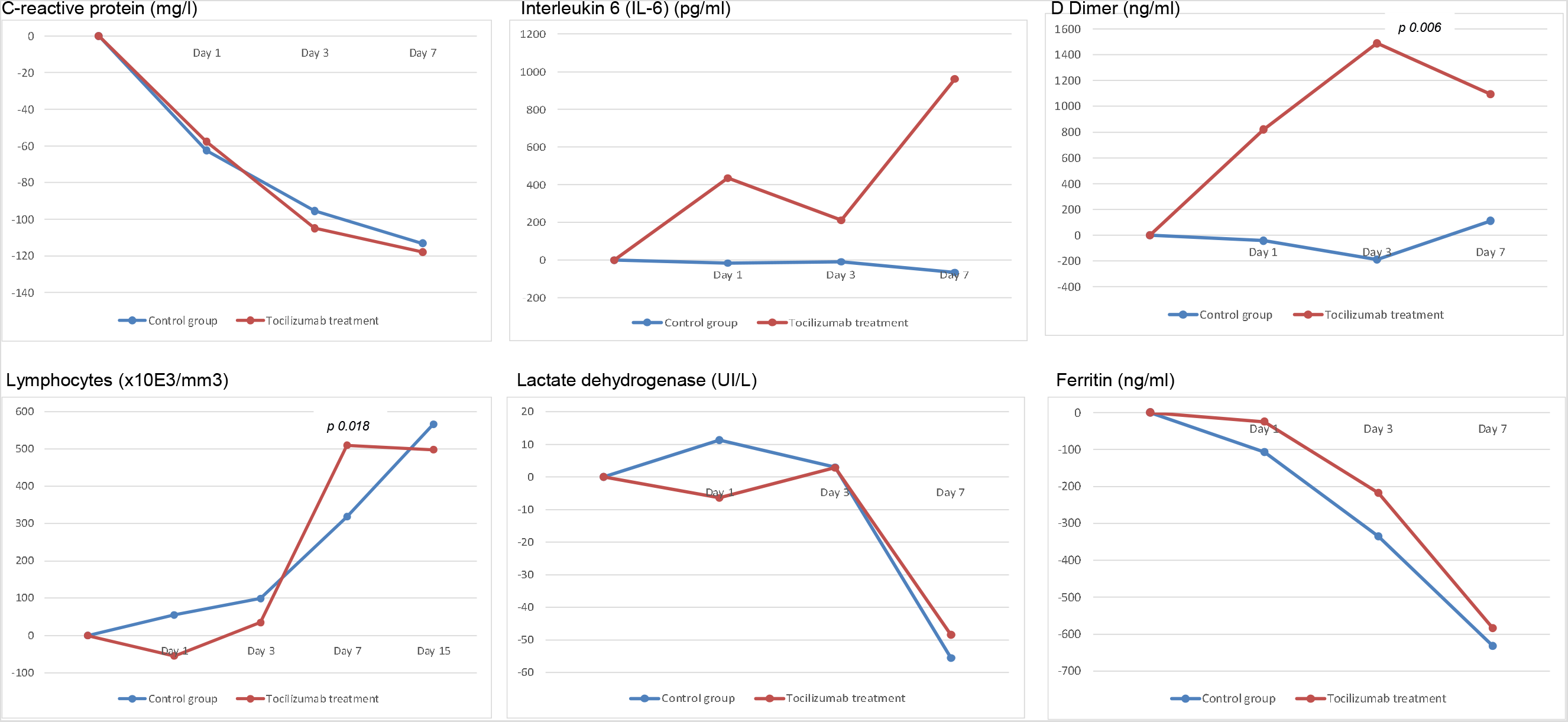
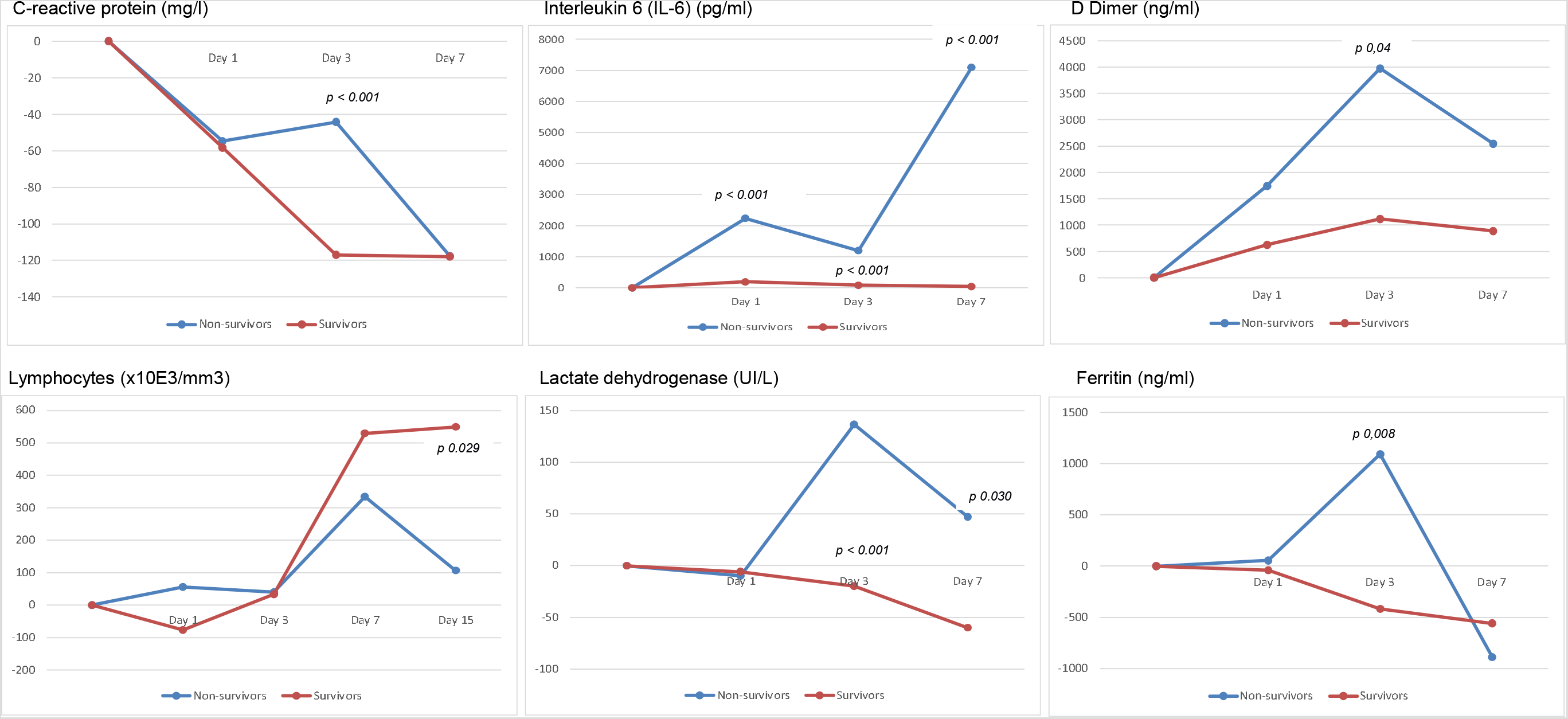
Variation over time of prognostic biomarkers

No differences in variation were observed according to tocilizumab exposure in biomarkers except for DD at D3 (higher in tocilizumab-treated cases) and lymphocyte counts at D7 (lower in tocilizumab-treated cases). However, among tocilizumab-treated patients, significant differences between survivors and non-survivors were observed at D3 in variation of CRP, IL6, LDH, DD and ferritin. Interestingly, by D7 biomarker levels had returned to baseline, except for of IL-6, that showed a striking difference at D7, and LDH. Lymphocyte count variation evolved differently, with a late significant difference with baseline at D15 between survivors and non-survivors.

#### Adverse events

Adverse events were monitored during the study period in the tocilizumab group. Although up to 32.6% in the tocilizumab group compared to 30.3 % in the control group had an increase in SGOT/AST above the upper limit of normal, there were no significant differences. A total of 11 (4.1%) patients had serious adverse reactions related to tocilizumab, including hepatotoxicity with increased liver enzymes (3 patients) or bilirubin (2 patients), thrombocytopenia (1 patient), catheter-related superficial thrombophlebitis (2 patients), diarrhea (1 patient), and headache and ocular phosphenes (1 patient). Bacteremia without a source after tocilizumab administration was documented in 1 patient (0.4%). Median follow-up time was 12 days (7-18 days).

### DISCUSSION

Our results show that survival of patients with severe COVID-19 is higher in patients treated with tocilizumab than in those not treated. In particular, mortality is improved when tocilizumab is used concomitantly or shortly followed by steroids (first 48h). These results contribute to the body of evidence supporting the use of tocilizumab in SARS-CoV-2 infection. Randomised controlled studies are needed to confirm these results and establish the potential place of tocilizumab in the treatment of COVID-19.

Our data are consistent with other recent series that report a reduction in risk of death in patients with severe COVID-19 treated with tocilizumab compared to standard of care. Results from randomised controlled trials have not yet been published. To our knowledge, only observational studies, some of them comparing tocilizumab use with a control group, are published[14], [15], [16].

Several anti-inflammatory and immunomodulatory therapies are being used in severe cases of COVID-19 who are purportedly in the inflammatory phase of the disease. In many cases, more than one drug is used, complicating further the interpretation of the results. Some of these therapies could boost the effect of the others. Recently, steroid treatment has shown promise as a life-saving therapy in patients with COVID-19 (Recovery trial: https://doi.org/10.1101/ 2020.06.22.20137273, [17, 18]). When analyzing the effect of other treatments, it is important to consider the combined use of steroid therapy.

Prior studies do not report specifically on the combined effect of tocilizumab and post-baseline steroids, although in most of them steroids have been used concomitantly with tocilizumab in a considerable proportion of cases (17% SOC y 30% tocilizumab [14]:), close to 100% in some cases [19],[20]. A single-centre Italian study analyses jointly data regarding the administration of both tocilizumab and steroids, together or separately, and suggests the need for evaluating combined use of these therapies ([21]). The moment of administration of steroids according to tocilizumab administration might be decisive. A small non-controlled Spanish series suggests that patients treated with prior or concomitant steroid treatment had a better survival than those with steroids added later [19], whereas a short observational single-centre study shows an improvement in mortality in patients treated with tocilizumab when salvage steroids are added in a median of 2.3 days[22]. In the present series, tocilizumab adds to steroids administered to non-intubated cases with severe COVID-19 during the first 48h hours of presenting with respiratory failure despite of oxygen therapy (Brescia-COVID respiratory severity scale 2 or worse).

Our findings concerning IL6 levels variations after tocilizumab treatment are consistent with those reported by Quartuccio et al [23], who report higher levels of IL-6 early after tocilizumab in non-survivors compared to survivors. Although it is known that following tocilizumab administration there is an IL6 increase due to saturation of the receptors[24], differences between survivors and non survivors are significant and could indicate a subset of patients who could benefit from further anti-IL6 therapy. Another promising approach is the study of genetic predictors of response to tocilizumab in COVID-19, which would allow to better select patients who would benefit most from tocilizumab therapy[25].

In agreement with Conrozier et al[26], we also observed differences between survivors and non-survivors in other biomarkers early after tocilizumab treatment, that, if confirmed in other studies, would help select subsequent therapies. As suggested by other authors [27], a persistent increase of ferritin or DD may indicate an escape of inflammation by other routes amenable to different treatments aiming IL1 (in the case of increased ferritin) or coagulation (in the case of DD).

Some authors suggest that the administration of tocilizumab is more effective in severe cases (PaO2/FiO2< 150)[14], while others report better results in cases with less oxygen requirements [20]. In the present series, we were not able to find a cut-off point for SaO2/FiO2 to ascertain which patients would do better under tocilizumab treatment. Our data differ from those of Martínez-Sanz et al. [15] who observed an association of tocilizumab effect with CRP levels above 150. In comparison, all the cases included in our work had severe COVID-19 and homogeneously high CRP levels, which may have prevented to find this association.

In this study, we showed that tocilizumab is effective to reduce mortality of severe COVID-19, and adds to steroids use. The major strength of this study was its multicentre nature, with a high number of patients included, together with the comparison with controls not treated with tocilizumab. The IPTW to adjust for indication bias adds to the strength of the results.

However, this is an observational study and, as such, has the inherent limitations of this kind of studies. Indication bias for steroid treatment was not adjusted for. It is possible that steroid treatment was not evenly administered across the study population. That could possibly explain the high mortality in the subset of patients who received steroids-48h, but not tocilizumab. IL6 levels were available only in a limited number of patients. IL6 receptor is the target of tocilizumab treatment, and some expert recommendations for tocilizumab use are based on IL6 levels. However, determination of IL6 was not available in all centres, and results were not timely, so that in the majority of cases it was not used to decide to initiate tocilizumab. Nevertheless, it is interesting to confirm that, in those with available levels, higher levels after tocilizumab administration seem to be predictive of poor outcome.

The short follow-up time prevents us to detect important adverse events such as reactivation of latent infections in patients treated with tocilizumab, and particularly in those treated with tocilizumab and steroids. Thus, safety concerns might have been underestimated. To overcome this problem, ongoing registries have been put in place to identify medium-term complications.

Although our cohort was limited to non-ICU patients, tocilizumab effect in critical patients who already require mechanical ventilation or ICU admission deserves investigation. One recent article by Somers et al[28] suggests that tocilizumab in ICU patients is also beneficial, although the risk of infection is high.

Multiple therapies for COVID-19 are currently under investigation. It is important to define the precise role of those that are effective, as COVID-19 may require different treatments at different stages or with different presentation patterns. Our findings contribute to establish the role of tocilizumab in patients with severe COVID-19 presenting with respiratory failure despite of oxygen therapy.

Additional studies, in particular randomized controlled studies, are needed to confirm these results and to figure out the characteristics of patients who benefit more from tocilizumab treatment. Similarly, the best moment to use it during the course of the disease remains to be determined. Prospective clinical trials with combined use of tocilizumab and steroids are warranted, with administration of steroids before, during and after tocilizumab, to determine the moment in which the combination is optimal. Longer follow-up is needed to ascertain potential infectious reactivations in patients treated with tocilizumab alone or in combination with steroids.

## Data Availability

Data sharing: After publication, the data will be made available to others on reasonable requests to the corresponding authors. A proposal with a detailed description of study objectives and statistical analysis plan will be needed for evaluation of the reasonability of requests. Additional materials might also be required during the process of evaluation. De-identified participant data will be provided after approval from the principal researchers of the Hospital Puerta de Hierro-Majadahonda.

## Acknowledgments

We thank the members of the TOCICOV study group for their contribution to the work.

## TOCICOV Study Group Collaborators

HOSPITAL DEL MAR- PARC DE SALUT MAR. X Fernández Sala, P. Díaz Pellicer, S. Grau Cerrato.

HOSPITAL UNIVERSITARIO DE PUERTO REAL. José Manuel Dodero Anillo, Alberto Romero Palacios.

HOSPITAL UNIVERSITARIO VIRGEN DEL ROCÍO. Elisa Cubiles, Mª Ángeles Lobo Acosta, Reyes Fresneda Gutierrez, Alicia Marían Candón, Sonsoles Salto Alejandre, Jose Miguel Cisneros Herreros, Elisa Cordero Matía, Carmen Infante Dominguez, Juan Carlos Crespo Rivas, Macarena López Verdugo.

HOSPITAL UNIVERSITARIO DE CANARIAS. Isabel Suarez Toste. Paloma Díaz Pérez.

HOSPITAL UNIVERSITARI DE BELLVITGE. Francesca Mitjavila Villero, Guillermo Suárez Cuartin, Carlota Gudiol Gonzalez, Adriana Iriarte Fuster, Mercè Gasa Galmes, Sandra Pérez, Dolores Rodríguez Cumplido.

HOSPITAL UNIVERSITARIO Nra Sra. CANDELARIA. Arístides de León Gil, Emilio J. Sanz

HOSPITAL GENERAL UNIVERSITARIO DE VALENCIA. Francisco Sanz-Herrero, Francesc Puchades, Pilar Ortega-García.

C.H. UNIV. DE SANTIAGO DE COMPOSTELA. José Antonio Díaz-Peromingo, Carlos Rodríguez-Moreno.

HOSPITAL UNIVERSITARIO CLÍNICO SAN CARLOS Daniel Lozano, Ana Terleira HOSPITAL UNIVERSITARIO TORRECÁRDENAS. María Estela González Castro, Sergio Ferra Murcia, Elena María Gázquez Aguilera.

HOSPITAL UNIVERSITARIO DE LA PRINCESA. Elena Pintos-Sánchez, Pablo Zubiaur, Elena Santos-Molina, Marcos Navares-Gómez, Gina Mejía Abril.

HOSPITAL CENTRAL DE LA DEFENSA GÓMEZ ULLA. Amelia García-Luque, Miguel Puerro-Vicente, María Jesús Sánchez Carrillo.

HOSPITAL UNIVERSITARIO RAMÓN Y CAJAL. M Ángeles Gálvez Múgica, Mónica Aguilar Jiménez, Cristina Sánchez Díaz.

HOSPITAL UNIVERSITARIO PUERTA DE HIERRO. Patricia Mills-Sánchez, Manuel Valle, Gustavo Centeno, Concepción Payares, Antonio F. Bermejo, Elena Diago, Rosa Malo, Juan Antonio Vargas Nuñez, Cristina Avendaño Solá, Elena Muñez, Antonio Ramos.

HOSPITAL UNIV. GERMANS TRIAS I PUJOL Ana María Barriocanal, Ana Pilar Pérez-Acevedoc, Melani Núñez-Montero.

HOSPITAL UNIVERSITARIO GREGORIO MARAÑON. María Olmedo, Sofia De la Villa.

HOSPITAL UNIVERSITARIO VIRGEN DE LA VICTORIA Enrique Nuño Álvarez, MI Lucena.

HOSPITAL COSTA DEL SOL. Josefa Andrea Aguilar García, José Javier García Alegría.

## Disclaimer

The results, discussion and conclusions are from the authors and do not necessarily represent the position of the Institution.

## Data sharing

After publication, the data will be made available to others on reasonable requests to the corresponding authors. A proposal with a detailed description of study objectives and statistical analysis plan will be needed for evaluation of the reasonability of requests. Additional materials might also be required during the process of evaluation. De-identified participant data will be provided after approval from the principal researchers of the Hospital Puerta de Hierro-Majadahonda.

## ABBREVIATIONS

95CI: 95% confidence interval
COPD: chronic obstructive pulmonary disease
COVID-19: coronavirus disease 2019
CoV: coronavirus
CRP: C- reactive protein
CT: computed tomography
HR: hazard ratio
ICU: Intensive care Unit
IL-6: interleukin-6
IPTW: inverse probability treatment weighting
IQR: interquartile range
LDH: Lactate dehydrogenase
PaO2/FiO2: arterial oxygen tension/inspiratory oxygen fraction
DD: D-dimer
RRR: relative risk reduction
SARS-CoV: severe acute respiratory syndrome coronavirus
SD: standard deviation
SGOT/AST: aspartate aminotransferase
SaO2: arterial oxygen saturation
Steroids: glucocorticoids or corticoids

---

**Appendix Table 1.**
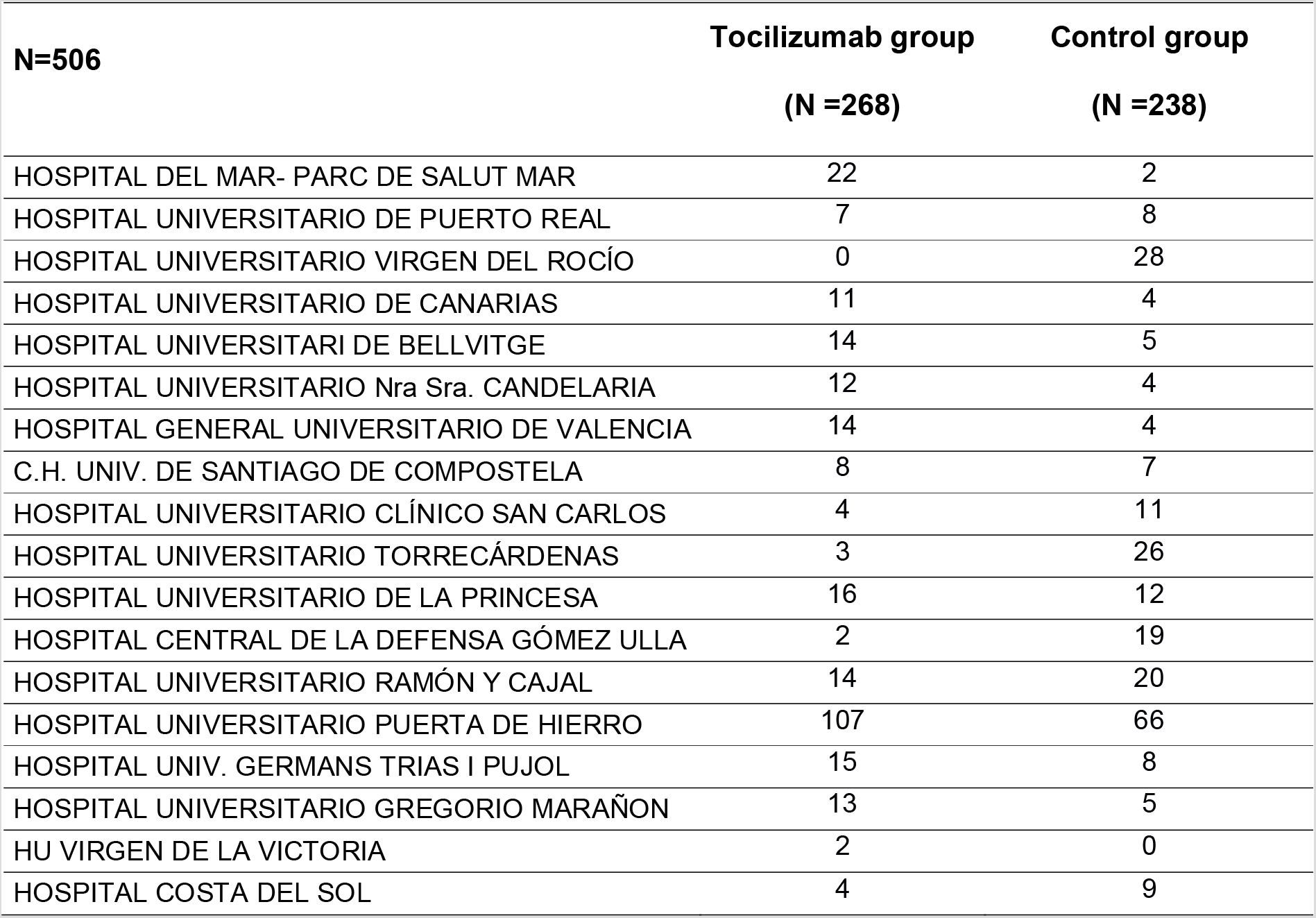
List of participating Centers and their contributions to the sample of 506 patients.

**Appendix Table 2.** Comparison between survivors and non-survivors.

| **N=506** | **Non-survivors** | **Survivors** | **p value** |
| --- | --- | --- | --- |
|  | **(N =120)** | **(N =386)** |  |
| Gender (Men), n (%) | 77 (64.2) | 247 (64.0) | >0.999 |
| Age, Mean (SD) | 78.8 (9.2) | 64.6 (12.6) | <0.001 |
| Underlying Medical Conditions, n (%) |  |  |  |
| High Blood pressure | 91 (75.8) | 184 (47.7) | <0.001 |
| Cardiovascular disease | 54 (45.0) | 85 (22.0) | 0.258 |
| Diabetes | 47 (39.2) | 99 (25) | 0.006 |
| Chronic kidney disease | 23 (19.2) | 30 (7.8) | 0.001 |
| Onco-hematologic | 10 (8.3) | 14 (3.6) | 0.047 |
| Chronic lung disease | 38 (31.7) | 57 (14.8) | <0.001 |
| Transplant (SOT/SCT) | 4 (3.3) | 6 (1.6) | 0.258 |
| Neurologic | 32 (26.7) | 25 (6.5) | <0.001 |
| Liver disease | 10 (8.3) | 12 (3.1) | 0.021 |
| HIV | 1 (0.8) | 4 (1) | >0.999 |
| NSAIDs | 10 (8.3) | 27 (7) | 0.688 |
| ACE inhibitors | 32 (26.7) | 61 (15.8) | 0.010 |
| ARBs | 34 (28.3) | 71 (18.4) | 0.028 |
| Treatment, n (%) |  |  |  |
| Hydroxychloroquine | 108 (90) | 374 (96.9) | 0.005 |
| Lopinavir/Ritonavir | 75 (62.5) | 329 (85.2) | <0.001 |
| Azithromycin | 67 (55.8) | 228 (59.1) | 0.596 |
| Remdesivir | 1 (0.8) | 1 (0.3) | 0.418 |
| Interferon | 31 (25.8) | 165 (42.7) | 0.001 |
| Steroids | 93 (77.5) | 299 (77.5) | >0.999 |
| - Steroids prior to D0 | 32 (26.7) | 81 (21.0) | 0.210 |
| - Steroids after D0 | 55 (53.4) | 215 (55.7) | 0.738 |
| WHO classification D0 |  |  |  |
| Admitted, no oxygen therapy | 1 (0.8) | 9 (2.3) | 0.054 |
| Admitted with oxygen therapy | 105 (87.5) | 355 (92) |  |
| High flow | 14 (11.7) | 22 (5.7) |  |
| Laboratory values, Mean (SD) |  |  |  |
| Lymphocytes (x10E3/mm<sup>3</sup>) | 755.1 (594.2) | 901.3 (498.1) | 0.008 |
| Lactate dehydrogenase (UI/L) | 502.1 (243.1) | 440.8 (275.7) | 0.029 |
| D Dimer (ng/ml) | 3029.5 (6921.4) | 1788.7 (2801.1) | 0.058 |
| C-reactive protein (mg/l) | 164.5 (101.2) | 143.9 (110.0) | 0.069 |
| Ferritin (ng/ml) | 1335.4 (1507.7) | 1597.1 (2344.5) | 0.384 |
| Interleukin 6 (IL-6) (pg/ml) | 294.5 (576.2) | 157.3 (382.2) | 0.059 |
| Chest CT (at hospital admission), n (%) |  |  | 0.154 |
| Normal imaging | 1 (0.9) | 12 (3.4) |  |
| Unilateral pneumonia | 15 (14.2) | 29 (8.1) |  |
| Bilateral interstitial pneumonia | 29 (27.4) | 95 (26.6) |  |
| Patchy bilateral pneumonia | 34 (32.1) | 143 (40.1) |  |
| Confluent bilateral pneumonia | 27 (25.5) | 78 (21.8) |  |
| Tocilizumab treatment, n (%) | 45 (37.5) | 223 (57.8) | <0.001 |
| Initial tocilizumab dose | 553.7 (115.7) | 528.4 (108.1) | 0.180 |
| Cumulative tocilizumab dose | 1406.7 (219.7) | 1526.7 (257.7) | 0.329 |
| Days from onset of symptoms to tocilizumab, Mean (SD) | 12.5 (6.3) | 11.5 (4.9) | 0.359 |
| Time from onset of symptoms to D0 | 9.2 (5.9) | 10.5 (5.0) | 0.017 |

**Appendix Table 3.** Characteristics of patients according to tocilizumab and steroid-48 exposure.

| **N=506** | **CONTROL Cohort**<br>**(N=151)** | **TOCILIZUMAB**<br>**only (N=149)** | **TOCILIZUMAB**<br>**+ STEROIDS-48**<br>**(N=119)** | **STEROIDS-48**<br>**only (N=87)** | **p value** |
| --- | --- | --- | --- | --- | --- |
| Gender (Men), n (%) | 59 (39.1) | 46 (30.9) | 38 (31.9) | 39 (44.8) | 0.106 |
| Age, Mean (SD) | 70.7 (14.2) | 65.9 (12.9) | 63.9 (10.0) | 72.4 (14.4) | 0.001 |
| Underlying Medical Conditions, n (%) |  |  |  |  |  |
| High Blood pressure | 91 (60.3) | 79 (53) | 51 (42.9) | 54 (62.1) | 0.014 |
| Cardiovascular disease | 44 (29.1) | 29 (19.5) | 36 (30.3) | 30 (34.5) | 0.055 |
| Diabetes | 34 (22.5) | 44 (29.5) | 34 (28.6) | 34 (39.1) | 0.060 |
| Chronic kidney disease | 20 (13.2) | 14 (9.4) | 9 (7.6) | 10 (11.5) | 0.459 |
| Onco-hematologic | 11 (7.3) | 7 (4.7) | 5 (4.2) | 1 (1.1) | 0.193 |
| COPD | 33 (21.9) | 23 (15.4) | 23 (19.3) | 16 (18.4) | 0.560 |
| Transplant (SOT/SCT) | 2 (1.3) | 4 (2.7) | 2 (1.7) | 2 (2.3) | 0.845 |
| Neurologic | 24 (15.9) | 10 (6.7) | 5 (4.2) | 18 (20.7) | <0.001 |
| Liver disease | 5 (3.3) | 4 (2.7) | 6 (5.0) | 7 (8.0) | 0.223 |
| HIV | 1 (0.7) | 2 (1.3) | 0 (0.0) | 2 (2.3) | 0.381 |
| NSAIDs | 17 (11.3) | 10 (6.7) | 9 (7.6) | 1 (1.1) | 0.038 |
| ACE inhibitors | 23 (15.2) | 29 (19.5) | 24 (20.2) | 17 (19.5) | 0.695 |
| ARBs | 34 (22.5) | 26 (17.4) | 23 (19.3) | 22 (25.3) | 0.474 |
| Treatment, n (%) |  |  |  |  |  |
| Hydroxychloroquine | 136 (90.1) | 144 (96.6) | 118 (99.2) | 84 (96.6) | 0.003 |
| Lopinavir/Ritonavir | 106 (70.2) | 133 (89.3) | 107 (89.9) | 58 (66.7) | <0.001 |
| Azithromycin | 77 (51.0) | 95 (63.8) | 68 (57.1) | 55 (63.2) | 0.108 |
| Remdesivir | 1 (0.7) | 1 (0.7) | 0 (0.0) | 0 (0.0) | 0.710 |
| Interferon | 55 (36.4) | 65 (43.6) | 59 (49.6) | 17 (19.5) | <0.001 |
| Steroids prior to D0 | 26 (17.2) | 87 (58.4) | 0 (0.0) | 0 (0.0) | <0.001 |
| WHO classification D0 |  |  |  |  |  |
| Admitted, no oxygen therapy | 2 (1.3) | 4 (2.7) | 1 (0.8) | 3 (3.4) | 0.312 |
| Admitted, oxygen therapy | 142 (94.0) | 136 (91.3) | 108 (90.8) | 74 (85.1) |  |
| High flow | 7 (4.6) | 9 (6.0) | 10 (8.4) | 10 (11.5) |  |
| Laboratory values, Mean (SD) |  |  |  |  |  |
| Lymphocytes (x10E3/mm<sup>3</sup>) | 908.9 (585.2) | 818.2 (540.7) | 854.7 (363.1) | 892.8 (576.7) | 0.414 |
| Lactate dehydrogenase (UI/L) | 433.6 (379.9) | 449.8 (204.5) | 453.5 (161.3) | 505.4 (251.5) | 0.318 |
| D Dimer (ng/ml) | 1972.3 (2457.6) | 2367.7 (6315.1) | 1990.5 (3388.4) | 1913.9 (2816.3) | 0.244 |
| C-reactive protein (mg/l) | 143.5 (119.8) | 136.4 (99.1) | 165.9 (111.8) | 155.9 (94.4) | 0.733 |
| Ferritin (ng/ml) | 1022.7 (1038.1) | 1747.2 (1486.8) | 1610.5 (1364.2) | 1721.3 (4078.5) | 0.170 |
| Interleukin 6 (IL-6) (pg/ml) | 69.2 (64.9) | 217.6 (618.4) | 194.7 (219.8) | 92.3 (126.4) | 0.260 |
| Chest CT (at hospital admission), n (%) |  |  |  |  |  |
| Normal imaging | 8 (5.8) | 3 (2.3) | 1 (0.9) | 1 (1.2) | 0.148 |
| Unilateral pneumonia | 17 (12.4) | 13 (2.3) | 5 (4.5) | 9 (11.1) |  |
| Bilateral interstitial pneumonia | 40 (29.2) | 34 (25.6) | 32 (28.6) | 18 (22.2) |  |
| Patchy bilateral pneumonia | 47 (34.3) | 56 (42.1) | 44 (39.3) | 30 (37.0) |  |
| Confluent bilateral pneumonia | 25 (18.2) | 27 (20.3) | 30 (26.8) | 23 (28.4) |  |
| Tocilizumab treatment, n (%) |  |  |  |  |  |
| Initial tocilizumab dose |  | 519.5 (112.2) | 547.9 (104.4) |  | 0.038 |
| Cumulative tocilizumab dose |  | 1444.4 (278.9) | 1528 (228.4) |  | 0.473 |
| Days from onset of symptoms to D0, Mean (SD) | 8.5 (4.8) | 12.6 (6.1) | 10.6 (3.6) | 8.3 (4.5) | <0.001 |
| ICU admission-IMV | 16 (10.6) | 32 (21.5) | 26 (21.8) | 7 (8.0) | 0.003 |
| Death | 40 (26.5) | 32 (21.5) | 13 (10.9) | 35 (40.2) | <0.001 |

\*prior to tocilizumab
COPD: chronic obstructive pulmonary disease; SOT: solid organ transplantation; SCT: stem cell transplantation; D0: day 0; SD: standard deviation; N/A: non-applicable; SaO2/FiO2: oxygen saturation /inspiratory oxygen fraction; ICU: intensive care unit; CT: computed tomography scan; IL-6: Interleukin 6.

